# Flooding and elevated prenatal depression in a climate-sensitive community in rural Bangladesh: a mixed methods study

**DOI:** 10.1101/2024.11.25.24317922

**Authors:** Suhi Hanif, Jannat-E-Tajreen Momo, Farjana Jahan, Liza Goldberg, Natalie Herbert, Afsana Yeamin, Abul Kasham Shoab, Reza Mostary Akhter, Sajal Kumar Roy, Gabriella Barratt Heitmann, Ayse Ercumen, Mahbub Rahman, Fahmida Tofail, Gabrielle Wong-Parodi, Jade Benjamin-Chung

## Abstract

**Background:** Prenatal depression can have lasting adverse impacts on child health. Little is known about the impact of floods on prenatal depression in low- and middle-income countries.

**Methods:** We conducted a cross-sectional survey of 881 pregnant women from September 24, 2023 to July 19, 2024 in riverine communities in rural Bangladesh. We recorded participant-reported flooding in the past 6 months, administered the Edinburgh Postnatal Depression Scale (EPDS), and obtained water level data and remote sensing data on distance to surface water. We fit generalized linear and log-linear models adjusting for month, wealth, education, age, and gestational age. We conducted 2 focus group discussions with 20 adult women.

**Findings:** 3.6% of compounds were flooded in the past 6 months. Compound flooding was associated with elevated depression (adjusted prevalence ratio (aPR) = 2.08, 95% CI 1.14, 3.51) and thoughts of self-harm (aPR=8.40, 95% CI 4.19, 16.10). Latrine flooding was associated with higher depression (aPR=3.58, 95% CI 1.49, 7.29)). Higher water levels and shorter distance to permanent surface water were significantly associated with mean EPDS scores. Focus groups revealed that domestic violence, inadequate sanitation, gendered vulnerabilities in accessing latrines, childcare difficulties, and food insecurity were key drivers of depression due to floods. Flood preparedness strategies included relocation, storing food, and home modifications.

**Interpretation:** Flooding, higher water levels, and proximity to water bodies were associated with prenatal depression in a rural, low-income setting. Inadequate sanitation and hygiene infrastructure were particularly strong drivers of depression.

**Funding:** Eunice Kennedy Shriver National Institute of Child Health and Human Development

**Research in Context:** *Evidence before this study:* We searched SCOPUS titles, abstracts, and keywords as follows: (antenatal OR prenatal OR perinatal OR prepartum OR pregnan* OR antepartum OR maternal) AND (depress* OR "mental health") AND (flood*). After filtering to include research articles focused on humans, we identified 35 articles, including 3 protocols, 4 reviews, 22 research articles in high-income settings, and 7 research articles in low- or middle-income countries (LMICs). In high-income settings, two studies have found that flooding is associated with prenatal depression. A review of the influence of extreme weather events on maternal health in LMICs only found one study that investigated the relationship between flooding and mental health (specifically coping) in pregnancy, but it did not measure depression. The search did not yield any studies that have investigated the relationship between flooding and prenatal depression in LMICs.

*Added value of this study:* To our knowledge, this is the first study estimating the association between flooding and prenatal depression in an LMIC. In a cross-sectional survey in a flood-prone region of rural Bangladesh, we found that flooding of the household compound and latrine was associated with higher prenatal depression prevalence. Proximity to surface water and higher water levels were associated with higher Edinburgh Postnatal Depression Scale scores. Our study describes flood preparedness strategies used by pregnant women and their households to inform climate adaptation in rural, riverine communities. We observed that these strategies largely focused on short-term mitigation rather than long-term resilience as more than half of all strategies involved temporary relocation. Additionally, we qualitatively examined gender-specific vulnerabilities related to flooding to understand mechanisms through which flooding may contribute to prenatal depression, with the aim of informing targeted flood resilience interventions for pregnant women. Women reported increased domestic violence during floods, gendered vulnerabilities in accessing latrines, and childcare difficulties, and food insecurity during floods.

*Implications of all the available evidence:* Our findings underscore the need to integrate maternal mental health care into climate resilience policy in flood-prone regions to prevent adverse downstream effects on child development and birth outcomes. Though pregnant women described multiple adaptation approaches, strong associations between flooding and depression indicate that existing adaptation methods fall short of supporting climate-related resilience. Climate-resilient water, sanitation, and hygiene (WASH) infrastructure may be particularly important to prenatal mental health during floods. Interventions may be most effective if targeted to women residing in areas closest to surface water.

## Introduction

Extreme weather events such as flooding, cyclones, hurricanes, and wildfires can have negative impacts on mental health outcomes.^1^ This is particularly the case in low- and middle-income countries (LMICs), which are more vulnerable to climate hazards. In South Asia, flooding is projected to increase under climate change, and it is associated with depression, anxiety, and post-traumatic stress disorder.^2^

No prior studies have investigated the impacts of flooding on prenatal mental health in LMICs,^3^ where 26% of women experience perinatal depression^4^. In high income settings, flooding is positively associated with perinatal depression^5^; children born to women who experienced natural disasters, including flooding, during pregnancy performed relatively poorly in elementary school^6^. Maternal depression is associated with adverse outcomes such as preterm births^7^, low birth weight, and impaired child development^8^. Exposure to stress and excess glucocorticoids in utero may result in epigenetic changes and immunologic, metabolic, neuroendocrine, and cognitive impacts associated with illnesses over the life course.^9^ It is essential to understand the impacts of flooding on the mental health of populations most vulnerable to climate change in order to inform climate adaptation strategies that promote women’s mental health, and in turn, healthy births, and child growth and development.

Flooding in LMICs could influence prenatal mental health through multiple pathways, including reduced agricultural productivity; lower income; damage to household infrastructure; lack of access to safe water, sanitation, and hygiene; displacement; trauma; and interruptions to prenatal or obstetric care.^2^ In some populations, gender norms require women to stay within the home unless accompanied by men and to keep their clothes dry can make the process of coping with floods more difficult for women, and their typical responsibility to serve as children’s primary caregivers can impede their efforts to adequately prepare for and remain safe during disasters.^10^ Similarly, women with children living in a cyclone prone region of Bangladesh were found to have a higher prevalence of depression after a natural disaster compared to women who did not have children^11^.

At the 2023 COP28 UN Climate Change Conference, countries agreed to a framework for the Global Goal on Adaptation with the goal of enhancing “adaptive capacity, strengthening resilience and reducing vulnerability to climate change.”^12^ The framework lays out areas that will require adaptation interventions, including health, but no measurable indicators of specific strategies have been identified, in part due to a need for more evidence. Recent reviews of adaptation strategies for mental health globally and for public health in LMICs both concluded that there is a dearth of evidence on this topic.^13^ For example, few studies have investigated individual- or household-level adaptation strategies to flooding in Bangladesh or other LMICs.^14^

We conducted a mixed methods study, including focus group discussions and a cross-sectional structured survey, to investigate the association between flooding and prenatal depression in a low-income, climate-vulnerable setting in rural Bangladesh. To generate evidence that will inform future adaptation interventions for climate resilience, we identified household strategies for preparing and responding to floods.

## Methods

### Study design

This analysis used data from the baseline survey of the Cement-based flooRs AnD chiLd hEalth (CRADLE) trial, an individually randomized trial in Sirajganj and Tangail districts, Bangladesh that will test whether replacing soil floors with concrete floors improves maternal and child health (NCT05372068).^15^ Households in our study were located in a flood-prone region adjacent to the Jamuna river in northeastern Bangladesh. The study population includes vulnerable communities living on riverine sandbars (locally known as chars), that are particularly at risk of flooding. The cross-sectional baseline survey was administered to pregnant women enrolled in the trial. The eligibility criteria for enrollment in the trial were as follows: households with a woman in her second or third trimester of pregnancy, household floor constructed of soil, household wall not made of earth, and no plan to relocate for 3 years. The survey included questions about flood history and flood preparation practices, a module on maternal depression, as well as household demographic characteristics, maternal education, assets, animal husbandry, and child illness.

### Exposures

We measured self-reported flooding with a recall period of 6 months. Participants were asked to report whether floods occurred within their union (the smallest rural administrative unit in the local government), their compound (typically 2-4 households of blood relatives that share latrines and food), or their latrine. Additionally, we obtained data on observed water levels in meters above mean sea level (mMSL) for Sirajganj District from the Bangladesh Water Development Board Flood Forecasting and Warning Centre. Water level was recorded daily at three-hour intervals by a designated gauge reader from the Bangladesh Water Development Board. We calculated the mean and maximum water levels across Sirajganj District in the 6 months prior to each participant’s survey date. We obtained data on seasonal and permanent surface water levels at 30 meter resolution from the Global Surface Water Explorer dataset^16^. We defined seasonal surface water as an area which is underwater for less than 12 months per year and permanent surface water as water that is present year-round. We calculated the distance from each household to the nearest pixel in which seasonal and permanent surface water was present as well as the proportion of the area around each household that contained seasonal and permanent surface water (radii: 10, 25, 50, 75, 100, 250, 500, 1000m) on the date of survey collection.

### Outcomes

We measured depressive symptoms among pregnant women using the Edinburgh Postnatal Depression Scale (EPDS). This tool has previously been validated for evaluating prenatal depression in rural South Asian settings. The EPDS score ranges from 0-30, with higher values indicating more severe depressive symptoms. We administered the validated Bengali version of the tool^17^. The instrument was administered by two enumerators with bachelor’s degrees who completed 7 days of training. There was an initial pilot with 25 pregnant women in the icddr,b hospital in Dhaka, followed by a second pilot in Chauhali sub-district. We classified women as moderately or severely depressed for total EPDS scores > 9.5 and as severely depressed for total EPDS scores > 13. To assess associations with individual EPDS questions, we created binary indicators. Responses that indicate no or few depressive symptoms were coded as 0 and responses that indicate depressive symptoms for that particular question were coded as 1.

### Statistical analyses

We pre-specified the statistical analysis plan (https://osf.io/vw9bj/), and we note deviations in Appendix 1. We estimated prevalence ratios for moderate or severe depression, severe depression, depressive symptoms for individual EPDS questions, and mean differences in the EPDS score using generalized linear models with a Gaussian family for the EPDS score and a binomial family for the prevalence of depression. We fit both crude and adjusted models. Potential confounders included the month of survey completion, a wealth index generated using the first principal component of household assets, women’s age in years, women’s years of education, their spouse’s years of education, and gestational age in weeks. We only adjusted for covariates associated with each outcome (likelihood ratio test p-value < 0.2). To assess potential residual unmeasured confounding, we calculated E-values for measures of association with flooding ^18^. The E-value is the minimum strength of association that must be present between an unmeasured confounder and both the exposure and the outcome to explain away the estimated measures of association^18^. Because the survey was cross-sectional and there were limited missing values, we performed a complete case analysis. Replication scripts are available here: https://github.com/jadebc/flood-depression-public

### Focus group discussions

To understand pathways through which flooding increased the risk of prenatal depression and to identify any further preparatory factors contributing to flooding resilience, we conducted focus group discussions (FGDs) with women not part of the CRADLE trial but who resided in Sirajganj and Tangail districts and were exposed to flooding risk. FGDs are a method of collecting qualitative data in a small group of participants, administered by a moderator, with prompts and discussion regarding participants’ lived experiences and perceptions. Participants were adult (age 18+) females residing in the CRADLE study region. Two FGDs were conducted in Bengali with 9 to 11 participants per group on August 13 and 14, 2024, and discussions lasted for up to one hour. A trained moderator conducted each FGD and audio-recorded the session, and other research staff took notes to aid in subsequent transcription and analysis. The moderator used a FGD guide developed by the research team in English before translation into Bengali. The guide began with introductions of participants before covering these topics in order: experiences with extreme weather, adaptation strategies, income impacts, displacement and shelters, migration, school enrollment and attendance, health, menstruation during floods, and anticipatory behaviors.

Audio recordings of the FGDs were transcribed into Bengali and then translated to English for analysis. The research team then coded employing a deductive approach of the English version of the transcripts using NVivo 14 based on the predetermined themes of flood preparedness and flood related vulnerabilities for women.

This study was approved by the International Centre for Diarrhoeal Disease Research, Bangladesh (icddr,b) Ethical Review Committee (PR-22069) and the Stanford Institutional Review Board (63990).

## Results

We administered our survey to 881 pregnant women, from September 24, 2023 to July 19, 2024. On average, women were 24.5 years old, 23.5 weeks pregnant and had 6.2 years of education (Appendix Table 1). More than half of the households (59%) had a monthly income less than USD 100 and the average household size was 5 members. The majority of homes were constructed with a tin roof and tin wall, and all homes had soil floors because having a soil floor was an enrollment criterion for the parent trial. All households had basic, improved water, 71% had unimproved sanitation, and 23% had a handwashing station with soap and water.

10.3% of households reported that their union was flooded for at least one day in the past six months (Table 1, Figure 1). In the past 6 months, the compound was flooded for at least one day in 3.6% (n) of households and the average length of flooding was 2.3 days. Three households (0.3%) reported that the inside of their home was flooded during this period and the flood lasted for an average of 14 days. Within the same recall period, flooding affected the tubewell (a type of well where a pipe is bored underground) in 0.6% of homes and the latrine in 1.2% of homes with the mean flooding duration being 12.5 days and 11.4 days, respectively. Rainfall primarily occurred between April and October (Figure 2a). The measured water level was highest between July and September (Figure 2b). During the recall period, women reported flooding events between the months of July and October (Figure 2b). Participant-reported flooding events tended to co-occur with elevated water levels. On average, the proportion of the 1km around each compound that contained permanent surface water was 0.014 (range: 0.000 to 0.199). The proportion of the 1km around each compound that contained seasonal surface water varied, with higher values during the rainy season and lower values in April and May, which tend to be hottest (Figure 2c).

**Figure 1.**
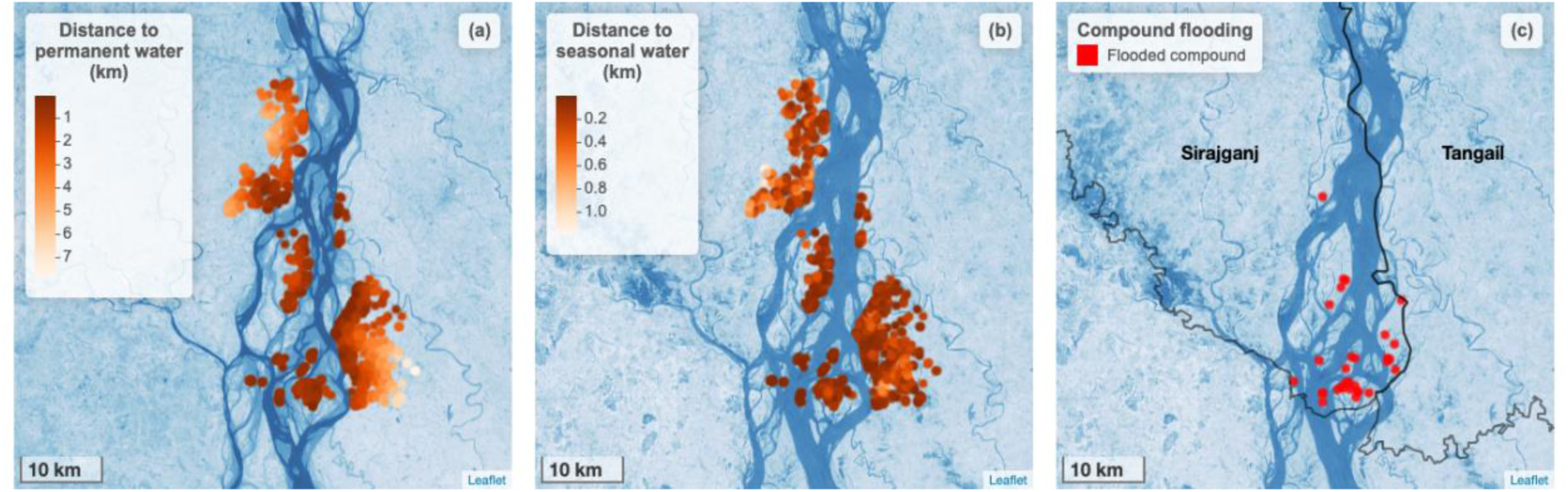
Surface water proximity and compound flooding in each study household. a) Points indicate the kilometers from each study household to permanent surface water. Base map shows permanent water bodies present during the dry season (November 2022 to May 2023). b) Points indicate the kilometers from each study household to seasonal surface water. Base map shows seasonal water bodies present during rainy season (June 2023 to October 2023). c) Red circles indicate households that experienced flooding in their compound in the 6 months prior to the study. Base map shows seasonal water bodies present during rainy season. In all panels, the base map is based on the Landsat 8-Day TOA Reflectance Composite. Darker shades of blue indicate a higher likelihood that surface water was present

**Figure 2.**
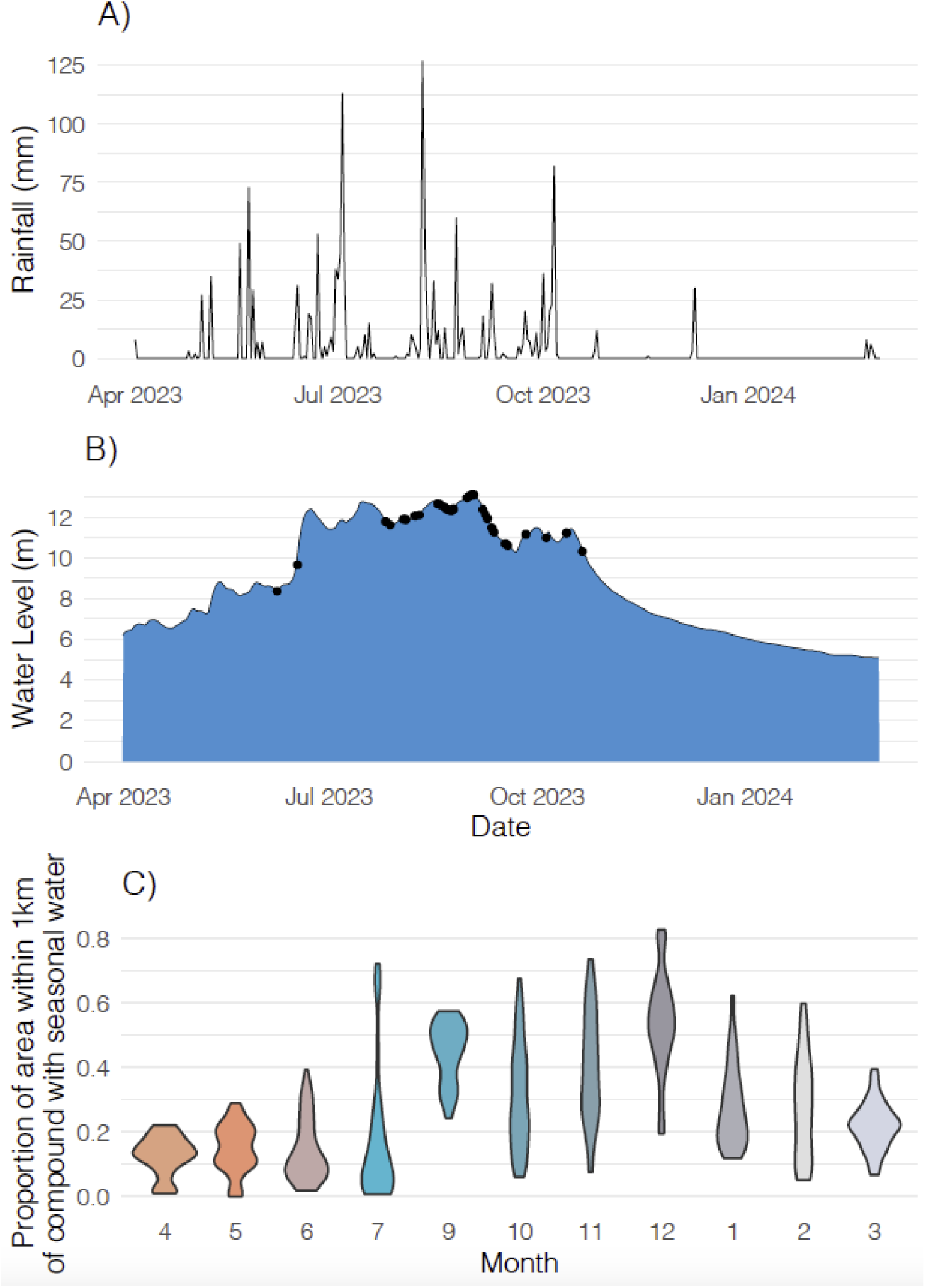
Rainfall, water level, and seasonal surface water patterns over time. a) Total daily mm of rainfall in Sirajganj district. b) Water level (meters above sea level) in Sirajganj district by date during the recall period for the study. Black points indicate dates when there was at least one flooding event reported by a participant. c) Violin plots showing the distribution of the proportion of area within 1km of compounds with seasonal surface water. The 1km radius is shown since all study households had seasonal surface water within this radius during the study period. Orange shades indicate the hottest months; teal shades indicate rainy season months; gray shades indicate drier and cooler months.

**Table 1:**
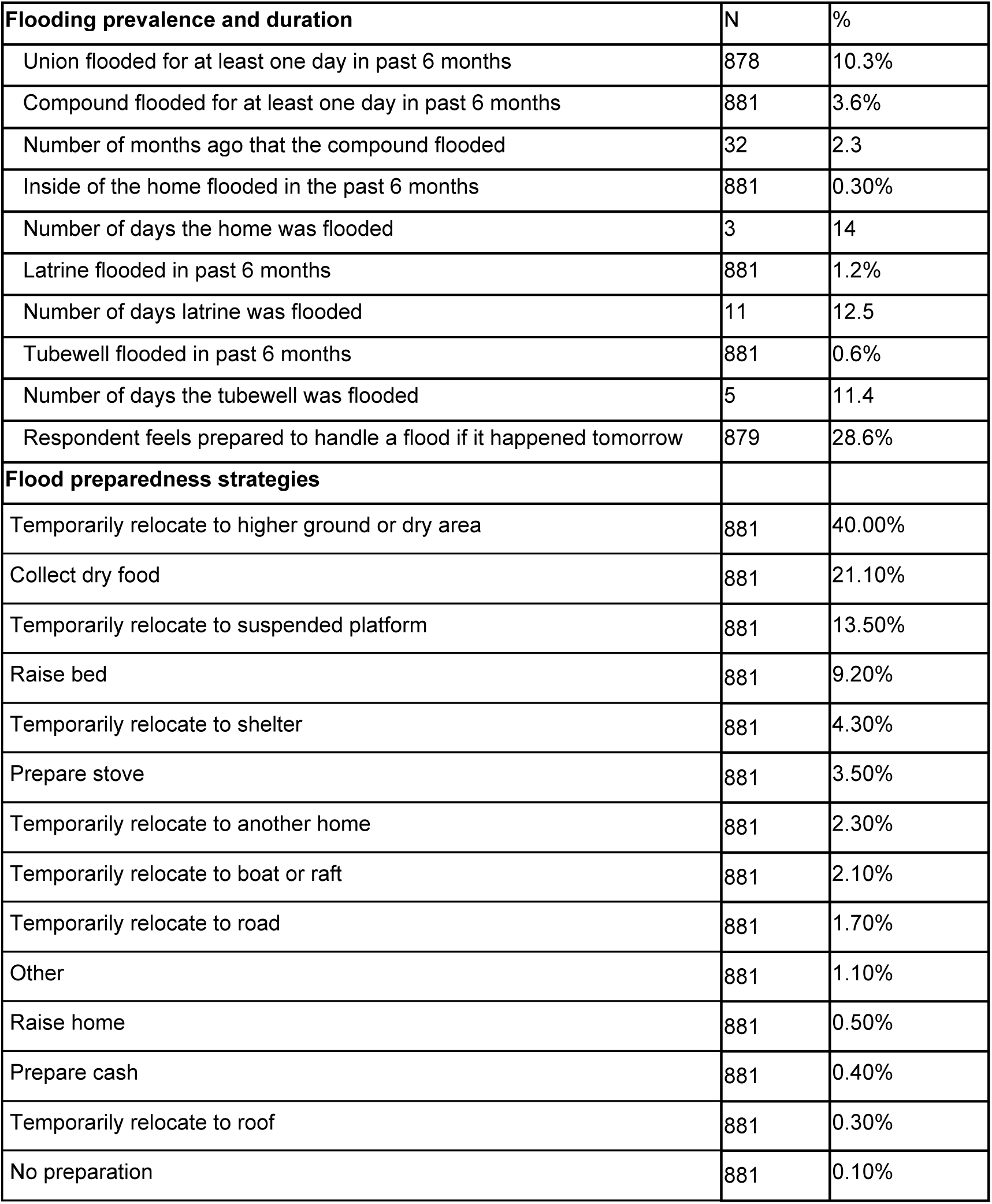
Prevalence of flooding and practices for flood preparedness.

Only 28.6% of women reported feeling prepared to handle a flood if it occurred the next day. One of the most common flood preparedness strategies was temporarily moving to a safe location (Table 1). Nearly half of respondents (40.0%) said they would move to higher ground or a dry area during a flood. Other temporary relocation strategies included living on a suspended platform (13.5%), moving to a flood shelter (4.3%), moving to someone else’s home (2.3%), living on a boat or raft (2.1%), and living on the road (1.7%). Common preparedness strategies other than relocation included collecting dry food (21.1%), preparing a portable stove (3.5%), and raising the height of the bed (9.2%).

Flooding was associated with higher prevalence of prenatal depression (Table 2). Women who experienced latrine flooding had an 3.58-fold (95% CI 1.49, 7.29) higher prevalence of depression and those who experienced compound flooding had a 2.08-fold (95% CI 1.14, 3.51) higher prevalence of depression compared to those who did not experience either type of flooding. Latrine flooding was also associated with a 3.58-fold (95% CI 1.49, 7.29) higher prevalence of depression and 4.33-fold (95% CI 1.01, 12.69) higher prevalence of severe depression compared to no latrine flooding. Flooding at the union level was associated with a 1.41-fold (95% CI 0.90, 2.11) higher prevalence of prenatal depression compared to no flooding in the union, but the confidence interval included the null. E-values for depression analyses indicated that unmeasured confounding would have had to be strong to fully explain estimated associations between compound or latrine flooding and depression (Appendix Table 2).

**Table 2:** Association between flooding and depression.

|  | N | Prevalence among exposed | Prevalence among unexposed | Crude prevalence ratio (95% CI) | Adjusted * prevalence ratio (95% CI) |
| --- | --- | --- | --- | --- | --- |
| **Moderate or severe depression** |  |  |  |  |  |
| Flooded latrine | 881 | 63.6% | 19.7% | 3.24 (1.38, 6.38) | 3.58 (1.49, 7.29) |
| Flooded compound | 881 | 43.8% | 19.3% | 2.26 (1.25, 3.76) | 2.08 (1.14, 3.51) |
| Flooded union | 878 | 27.8% | 19.3% | 1.44 (0.92, 2.16) | 1.41 (0.90, 2.11) |
| **Severe depression** |  |  |  |  |  |
| Flooded latrine | 881 | 27.3% | 7.2% | 3.77 (0.92, 10.14) | 4.33 (1.01, 12.69) |
| Flooded compound | 881 | 12.5% | 7.3% | 1.71 (0.52, 4.15) | 1.42 (0.42, 3.54) |
| Flooded union | 878 | 8.9% | 7.2% | 1.23 (0.54, 2.43) | 1.19 (0.52, 2.37) |

\*Adjusted models adjusted for any of the following covariates that were associated with each outcome (likelihood ratio p-value<0.2): month, wealth index, mother's years of education, spouse's years of education, mother's age, gestational age

On average, women who experienced latrine flooding had an EPDS score that was 3.87 points (95% CI 1.14, 6.60) higher compared to those without latrine flooding (Appendix Table 3). Women who experienced compound flooding had a 2.34 point (95% CI 0.71, 3.96) higher EPDS score than those who did not experience compound flooding. We did not find evidence of an association between union flooding and the EPDS score.

We also measured the association between individual EPDS questions and compound flooding (Table 3). Women who experienced compound flooding had a 2.06-fold (95% CI 1.15, 3.41) higher prevalence of being unable to see the funny side of things and a 1.88-fold (95% CI 1.03, 3.18) higher prevalence of finding it difficult to look forward to enjoyment compared to women who did not experience compound flooding. Additionally, compound flooding was associated with a 8.40-fold (95% CI 4.19, 16.10) higher prevalence of experiencing thoughts of self-harm. Other individual EPDS responses were not associated with flooding.

**Table 3:** Adjusted association between individual EPDS questions and compound flooding.

|  | Prevalence among exposed | Prevalence among unexposed | Adjusted prevalence ratio (95% CI) |
| --- | --- | --- | --- |
| Difficult to see the funny side of things | 46.9% | 20.3% | 2.06 (1.15, 3.41) |
| Difficult to look forward to enjoyment | 43.8% | 20.0% | 1.88 (1.03, 3.18) |
| Blamed self unnecessarily when things went wrong | 9.4% | 17.1% | 0.50 (0.12, 1.32) |
| Felt anxious or worried for no good reason | 21.9% | 19.9% | 1.09 (0.46, 2.18) |
| Felt scared or panicky for no good reason | 25.0% | 10.5% | 2.37 (1.05, 4.67) |
| Things getting on top of self | 31.2% | 22.6% | 1.26 (0.62, 2.29) |
| Difficulty sleeping due to unhappiness | 15.6% | 19.0% | 0.72 (0.26, 1.60) |
| Felt sad or miserable | 25.0% | 21.8% | 1.02 (0.46, 1.95) |
| Cried due to unhappiness | 28.1% | 13.9% | 1.84 (0.86, 3.47) |
| Thoughts of self-harm | 43.8% | 4.4% | 8.40 (4.19, 16.10) |

\*Adjusted models adjusted for any of the following covariates that were associated with each outcome (likelihood ratio p-value<0.2): for month, wealth index, mother's years of education, spouse's years of education, mother's age, gestational age

We did not observe associations between distance to any surface water and the prevalence of prenatal depression (Appendix Table 4). Each additional meter away from permanent surface water was associated with a 0.27 (95% CI 0.05, 0.49) lower mean EPDS score; we did not observe an association with seasonal surface water (Appendix Table 5). Overall, we did not observe associations between the proportion of the area near households that contained surface water and depression (Appendix Table 6).

There was a positive association between water level and mean EPDS score in adjusted models. A one meter increase in mean water level above sea level in the past 6 months was associated with 0.27 (95% CI 0.11, 0.42) higher mean EPDS score. We did not observe associations between water level and the prevalence of depression (adjusted PR = 1.03 (95% CI 0.95, 1.11)) or severe depression (adjusted PR = 1.07 (95% CI 0.94, 1.22)).

FGDs revealed numerous flooding-related vulnerabilities and adaptation strategies including housing security; food insecurity; income, savings, and livelihoods; health and physical safety; water, sanitation, and hygiene (Boxes 1-2). For most domains, participants reported more vulnerabilities than adaptation strategies; however, for housing security, participants mentioned several strategies for altering their homes during flooding events to reduce the chance of displacement due to damage to their homes or belongings. The number of discrete vulnerabilities mentioned was largest for water, sanitation, and hygiene, with women facing multiple challenges to bathing, maintaining menstrual hygiene, and relieving themselves during floods. While participants mentioned upgrades to their household structure to prepare for floods, none mentioned preparatory upgrades to their latrines.

### Box 1. Flooding-related vulnerabilities and adaptation strategies identified in focus groups with adult men and women

| Flooding-related vulnerabilities | Flooding adaptation strategies |
| --- | --- |
| **<u>Housing security</u>**<br>• Displacement<br>• Damage to home and belongings<br>**<u>Food security</u>**<br>• Damaged food, unable to travel to market to purchase food, especially for children<br>• Cannot operate stove to cook<br>• Dietary changes due to limited food availability<br>• Damage to food crops for household food consumption and income generation<br>**<u>Income, savings, and livelihoods</u>**<br>• No savings available to prepare for a flood<br>• Livestock/poultry drowning impacts women's livelihoods<br>• Going into debt due to borrowing for home preparation or purchasing extra food prior to floods<br>**<u>Health and physical safety</u>**<br>• Disease (e.g., skin rash, diarrhea)<br>• Venomous snakes entering the home<br>• Increased risk of drowning<br>• Domestic violence<br>• Harassment of girls in shelters<br>**<u>Water, sanitation, and hygiene</u>**<br>• Limited drinking water access<br>• Open defecation<br>• Using temporary, unhygienic latrine<br>• Relieving oneself less often<br>• Lack of privacy when using latrines<br>• Wading through flood water to access latrines<br>• Waiting in long queue to use neighbor's latrine<br>• Soiling oneself while waiting to access a latrine<br>• Difficult to bathe<br>• Difficult to maintain menstrual hygiene | **<u>Housing security</u>**<br>• Temporarily living on a suspended platform or on a roof<br>• Reinforce home structure (banana trees, bamboo poles, wrap foundation in polythene to prevent erosion)<br>• Raising the foundation of the home<br>**<u>Food security</u>**<br>• Storing food<br>• Using or borrowing a boat (e.g., to travel to buy food)<br>**<u>Income, savings, and livelihoods</u>**<br>• Saving money to prepare for a flood<br>• Moving livestock to raised ground or raised platform<br>**<u>Health and physical safety</u>**<br>• Raising beds (e.g., to keep children away from flood water)<br>**<u>Water, sanitation, hygiene</u>**<br>• Treating drinking water<br>**<u>Access to information</u>**<br>• Receiving flood warning text messages from non-governmental organizations |

### Box 2. Quotes from focus group discussion participants on how flooding influences their lives

*“Women cannot defecate anywhere (during floods). They need privacy. When a woman goes to someone else’s toilet, the surrounding people tease them. We feel shy so we go to other people’s houses secretly to use their toilet. During a flood, our dresses get wet while going to someone else’s home. There are long queues in every home to use the toilet. So, we move from house to house to find an unoccupied washroom. Sometimes we cannot control the pressure for such a long time and our dresses get spoiled due to it.”*

*“When the water level increases, we cannot eat properly to satisfy our hunger because we know that if we eat enough food to satisfy our hunger, we will have to use the washroom frequently. That is why we try to consume less. During the daytime, we go to a house which is not flooded and use their washroom. We have to go there by crossing flood water. Our clothes get wet and after returning home, we have to change it. That is why we try to consume less so that we do not have to use the washroom more than once a day.”*

*“We cannot have three meals a day. It is difficult to have at least one meal a day during floods. Even if we can manage one meal a day, we have to give most of the food to our children. Because if we eat according to our hunger, we will not be able to give them enough food. That’s why we give them enough food by sacrificing from our sides.”*

*“It is the month of floods. There is no income in the family. So, when I ask for something he (my husband) starts to argue with me. If I tell him to buy some groceries, he starts to hit me. So, I do not express myself, usually out of fear of violence. Still, sometimes I have to tell him when there is no way.”*

## Discussion

Flooding of compounds and latrines was associated with higher prevalence of prenatal depression and thoughts of self-harm in rural riverine communities in Bangladesh. Latrine flooding had a particularly strong association with prenatal depression. Higher water levels and shorter distances to permanent surface water were associated with higher EPDS scores. Focus group discussions substantiated these findings and revealed the multitude of ways in which flooding events influenced mental health by, for example, preventing pregnant women from bathing and relieving themselves. We also identified multiple ways that some women prepared for and/or remained resilient during flooding events, including home modifications, storing food, saving money, and treating drinking water.

Overall, the frequency of flooding and prevalence of depression in our study population was within the range of estimates in other studies in rural Bangladesh.^19^ Compared to a prior study in Bangladesh investigating flooding and depression in the general population, we found stronger associations with prenatal depression.^20^ This is consistent with our expectation given that women in Bangladesh are more negatively impacted by flooding than men^21^. Pregnant women could be more vulnerable to flooding because normal changes to the brain during pregnancy could interrupt necessary responses to disasters, and disasters could directly result in brain rewiring during pregnancy.^9^ Other studies have found that the severity of a natural disaster in pregnancy correlates with the intensity of perinatal depression.^9^ We found higher levels of suicidal ideation following floods than a prior study did in a flood-prone area of Bangladesh;^22^ this difference could reflect an acute response to a flooding event as opposed to a persistent flood risk.

The adaptation strategies households used to prepare for and respond to floods were similar to those reported in other studies in Bangladesh. A study conducted in remote chars, or sandbars, identified adaptation strategies such as treating drinking water, storing food, finishing work early in the morning, selling assets, and taking out loans.^14^ Additional flooding adaptation practices in Bangladesh include floating agriculture and raised houses, tubewells, and latrines.^23^ However, overall, adaptive capacity remains low: it is estimated that one third of households in northern Bangladesh have the capacity to recover from floods through adaptations such as altered agricultural practices.^24^ At the country level, Bangladesh ranks low in adaptation, with an ND-GAIN score of 27 in 2022 (maximum score 100) for its readiness to effectively invest in climate adaptations.^25^ Our findings suggest that the adaptation strategies in use in this population may be insufficient for climate resilience.

Our quantitative and qualitative analyses pointed to inadequate sanitation and hygiene during floods as a strong driver of prenatal depression. Women mentioned multiple vulnerabilities related to sanitation and hygiene but did not report any adaptation strategies (Box 1). In contrast, participants identified both vulnerabilities and multiple adaptation strategies for securing and reinforcing their home. A prior review identified multiple physical, financial, and social stressors resulting from a lack of access to sanitation, with stronger psychosocial impacts on women.^26^ Safe sanitation and hygiene is essential to women of childbearing age to maintain hygiene during menstruation, spotting during pregnancy, and bleeding for 4-6 weeks following miscarriage and postpartum.^27^ Our focus group findings were consistent with reports in prior studies that women experience shame and risk of violence when they must go outside the home setting to access safe water or sanitation, especially if their clothing is wet.^10^ Upgrading water, sanitation, and hygiene infrastructure to be climate resilient may be necessary to sustain women’s mental health during floods.

Other pathways through which flooding influenced women’s mental health included displacement, food insecurity, threats to physical safety, additional responsibilities in the home, and economic hardship, consistent with prior studies. Prior to floods, women in rural Bangladesh are traditionally responsible for preparing portable stoves and raising the height of the home or bed^28^; following floods, it may be more difficult to keep up with household responsibilities due to damaged or lost household assets.^10^ Women reported that they were less likely to eat during floods, consistent with prior studies.^21^ Major floods are associated with food insecurity in Bangladesh, with women who experienced a major flood having higher odds of depression even 2.5 years after the event^29^. Gender norms, including purdah, which requires women to remain within the compound unless accompanied by a male relative, can also prevent women from taking actions to prepare for or safely relocate during floods.^10^ Climate-resilient household construction that prevents flood water from entering the home and latrine and reduces the risk of temporary displacement may help reduce gender inequities in mental health following climate disasters.

A strength of our study is our mixed methods approach, which allowed us to quantify associations with flooding and to capture contextual and cultural factors contributing to depression following floods, consistent with recommended practices for research on climate and health.^30^ This study was subject to limitations. Estimates may be subject to residual confounding since we did not have data on potential confounders such as pre-pregnancy depression, antenatal care, pregnancy symptoms, domestic violence, or social support. However, our E-value analyses showed that confounders would have had to be very strong to explain away associations with compound and latrine flooding. Additionally, self-reported flooding may be subject to recall error or courtesy bias; however, a strength of our study is that we also included objectively measured exposures (distance to surface water, water level); scientific inferences across exposures were similar for analyses of depression scores. This internal consistency between analyses with different exposures supports the validity of our inferences.

In conclusion, our findings highlight the need to prioritize prenatal depression as a significant public health concern in flood-prone regions of Bangladesh, especially in the context of increasing climate-related hazards. Though study participants report multiple adaptation strategies, strong associations with depression suggest that current strategies are insufficient for climate resilience. Integrating depression screening into antenatal care services in flood-prone regions and among women who reside close to surface water, building flood-resilient homes and latrines, and post-disaster mental health support may improve mental health of pregnant women living in vulnerable areas.

## Data Availability

Data will be made public at the time of publication.

## Author contributions

SH, FJ, RMA, FT, LG, NH, GWP, MR, and JBC conceptualized and developed the study and its design. GWP, LG, NH, AY, AKS, FJ, RMA, and JM contributed to data collection. All authors contributed to the statistical pre-analysis plan. JM transcribed focus group discussions, and SH coded them. SH and JBC conducted the statistical analysis and wrote the first draft of the manuscript. All authors reviewed and approved the manuscript’s final draft.

## Disclaimer

The content of this manuscript is solely the responsibility of the authors and does not necessarily represent the official views of the National Institutes of Health.

## Appendix

**Appendix Table 1:** Characteristics and flood experiences of participants. * Sanitation and water defined using the WHO/JMP water and sanitation ladders.

| Characteristic | N | Mean/% |
| --- | --- | --- |
| **Participant characteristics** |  |  |
| Mother's years of education | 881 | 6.2 |
| Spouse's years of education | 881 | 5.2 |
| Mother's age in years | 881 | 24.5 |
| Gestational age in weeks | 881 | 23.5 |
| **Household characteristics** |  |  |
| Number of household members | 881 | 4.9 |
| Roof material is bamboo | 881 | 0.10% |
| Roof material is tin | 881 | 99.9% |
| Wall material is jute, bamboo, or straw | 881 | 3.5% |
| Wall material is brick or cement | 881 | 0.50% |
| Wall material is tin | 881 | 96% |
| Floor material is mud | 881 | 100% |
| Sanitation* |  |  |
| Basic | 881 | 17% |
| Limited | 881 | 12% |
| Unimproved | 881 | 71% |
| Open defecation of children <3 years | 881 | 21% |
| Handwashing station with soap and water | 881 | 23% |
| Basic water* | 881 | 100% |
| **Sociodemographic characteristics** |  |  |
| Monthly income < 12,001 taka | 881 | 59% |
| Has electricity | 881 | 97% |
| Owns mobile | 881 | 98% |
| Owns livestock | 881 | 79% |
| Father works in agriculture | 881 | 19% |
| **Flooding** |  |  |
| Union flooded for at least one day in past 6 months | 878 | 10.3% |
| Compound flooded for at least one day in past 6 months | 881 | 3.6% |
| Number of months ago that the compound flooded | 32 | 2.3 |
| Inside of the home flooded in the past 6 months | 881 | 0.30% |
| Number of days the home was flooded | 3 | 14 |
| Latrine flooded in past 6 months | 881 | 1.2% |
| Number of days latrine was flooded | 11 | 12.5 |
| Tubewell flooded in past 6 months | 881 | 0.6% |
| Number of days the tubewell was flooded | 5 | 11.4 |
| Respondent feels prepared to handle a flood if it happened tomorrow | 879 | 28.6% |

**Appendix Table 2:** E-values for prevalence ratios with depression.

| Outcome | E-value |
| --- | --- |
| **Moderate or severe depression** |  |
| Flooded latrine | 6.61 |
| Flooded compound | 3.58 |
| Flooded union | 2.16 |
| **Severe depression** |  |
| Flooded latrine | 8.13 |
| Flooded compound | 2.19 |
| Flooded union | 1.67 |

**Appendix Table 3:** Association between flooding and EPDS score.

|  | N | Mean among exposed | Mean among unexposed | Crude mean difference (95% CI) | Adjusted * mean difference (95% CI) |
| --- | --- | --- | --- | --- | --- |
| Flooded latrine | 881 | 9.5 | 5.7 | 3.76 (0.99, 6.53) | 3.87 (1.14, 6.60) |
| Flooded compound | 881 | 8.3 | 5.6 | 2.67 (1.03, 4.31) | 2.34 (0.71, 3.96) |
| Flooded union | 878 | 6.6 | 5.6 | 0.98 (-0.04, 2.00) | 0.86 (-0.14, 1.86) |

\*Adjusted models adjusted for any of the following covariates that were associated with each outcome (likelihood ratio p-value<0.2): for month, wealth index, mother's years of education, spouse's years of education, mother's age, gestational age

**Appendix Table 4.** Association between distance to surface water and moderate/severe depression and severe depression.

|  | N | Crude prevalence ratio (95% CI) | Adjusted * prevalence ratio (95% CI) |
| --- | --- | --- | --- |
| **Moderate or severe depression** |  |  |  |
| Seasonal water | 881 | 0.92 (0.29, 2.68) | 1.02 (0.32, 3.06) |
| Permanent water | 881 | 0.92 (0.81, 1.03) | 0.95 (0.83, 1.07) |
| **Severe depression** |  |  |  |
| Seasonal water | 881 | 0.13 (0.01, 0.91) | 0.19 (0.02, 1.37) |
| Permanent water | 881 | 0.79 (0.62, 0.96) | 0.86 (0.67, 1.06) |

\*Adjusted for month, wealth index, mother's years of education, spouse's years of education, mother's age, gestational age

**Appendix Table 5.** Association between distance to surface water and EPDS score.

|  | N | Crude mean difference (95% CI) | Adjusted * mean difference (95% CI) |
| --- | --- | --- | --- |
| Seasonal water | 881 | -1.67 (-3.72, 0.38) | -1.42 (-3.45, 0.62) |
| Permanent water | 881 | -0.35 (-0.56, -0.13) | -0.27 (-0.49, -0.05) |

\*Adjusted for month, wealth index, mother's years of education, spouse's years of education, mother's age, gestational age

